# Agentic Trial Emulation to Learn Health System-specific Drug Effects At Scale

**DOI:** 10.64898/2026.02.19.26346539

**Authors:** Justin Kauffman, Lisa Duan, Samuel Gelman, Eyal Klang, Ankit Sakhuja, Deepak L. Bhatt, Vivek Y. Reddy, Alexander W. Charney, Girish N. Nadkarni, Yuanhao Qu, Kexin Huang, Joshua Lampert, Benjamin S. Glicksberg

## Abstract

**Objective:** Electronic Health Record (EHR)-based trial emulation can support translation of randomized clinical trial (RCT) evidence into practice, yet emulations often diverge from published RCT results. We hypothesized that these discrepancies are structured and learnable properties of a health system’s data-generating process, and that autonomous agentic workflows can generate discrepancies at the scale required for cumulative learning.

**Materials and Methods:** We developed an agentic trial emulation framework that (1) uses an autonomous LLM agent (Biomni) to execute an end-to-end, instruction-driven emulation pipeline against an OMOP CDM database and (2) calibrates EHR estimates to RCT results with a Bayesian hierarchical model. Biomni performed protocol parsing, OMOP concept set construction, cohort building, confounder adjustment, and treatment effect estimation; it also synthesized literature-derived, comparison-specific priors for expected EHR–RCT disagreement. Five atrial fibrillation anticoagulation trials were emulated using Mount Sinai’s OMOP-mapped EHR, with three independent runs per trial to quantify agent-induced analytic variability. Discrepancies between EHR-derived and published log–hazard ratios were modeled as the sum of a literature-informed reproducibility expectation, an institution-specific systematic shift, and residual heterogeneity. Performance was assessed using leave-one-out cross-validation across four in-domain DOAC-versus-warfarin trials, with one out-of-distribution evaluation (apixaban versus aspirin).

**Results:** In pooled leave-one-out validation, calibration reduced mean absolute error from 0.567 to 0.224 log–hazard ratio (60.5% reduction) and achieved 100% empirical coverage of 95% posterior predictive intervals across held-out trials (4/4). The posterior institution-specific shift was consistently positive across folds (median 0.364–0.580), indicating systematic attenuation of DOAC benefit in the local EHR beyond literature-expected disagreement; residual heterogeneity was moderate (median 0.199–0.264). For the out-of-distribution AVERROES trial, calibrated error decreased from 0.379 to 0.051 (86.5% reduction), with the published effect within the 95% credible interval.

**Discussion and Conclusion:** Autonomous emulation with agents enables repeated, standardized trial replications that convert EHR–RCT disagreement into data for learning institution-level transport properties. Separating comparison-specific reproducibility expectations from system-level shifts yields calibrated, uncertainty-aware local interpretations of trial evidence.

## Introduction

Randomized clinical trials (RCTs) provide the strongest evidence for treatment efficacy, yet translating trial findings into decisions for a specific health system remains difficult [15,16]. Differences in patient mix, prescribing and adherence, co-interventions, outcome ascertainment, and coding practices can meaningfully alter how a trial-reported effect manifests in routine care [16,19]. Real-world evidence (RWE) derived from electronic health records (EHRs) is often used to bridge this gap, including through “target trial” emulation of RCT protocols [6]. However, EHR emulations frequently diverge from published RCT results, creating an interpretability problem [7,8,10,11]. Specifically, when estimates disagree, it is not clear if the discrepancy should be treated as evidence of bias, evidence of non-transportability, or both.

Most trial emulation workflows implicitly resolve this dilemma by treating the RCT result as ontologically privileged and interpreting EHR–RCT disagreement as methodological failure-residual confounding, incomplete protocol capture, measurement error, or data quality limitations [7,23,24]. Under this framing, concordance is accepted without further analysis and discordance is discounted. This approach discards information: discrepancies are not purely random artifacts, but arise from structured differences between randomized and observational data-generating processes [8,10,12,13] that depend on the drug comparison, endpoint definition, confounding architecture, and the local clinical and administrative context in which data are produced. In other words, disagreement between RCT and EHR estimates can encode how a health system systematically transforms trial evidence as it is expressed through routine care.

Learning from this structure requires scale. A single emulation provides limited insight, because any discrepancy can be attributed to idiosyncratic implementation choices or chance. By contrast, a systematic program of emulation across drugs, endpoints, and clinical contexts can accumulate a pattern of discrepancies that characterizes an institution’s “transport relationship” to external trial evidence, such as identifying when RWE is likely to align with RCT findings, when it is likely to deviate in a consistent direction, and where uncertainty remains irreducible. The core barrier is practical. End-to-end trial emulation remains labor intensive, requiring protocol interpretation, phenotype construction, cohort assembly, confounder adjustment, survival modeling, and extensive diagnostics [6,48,49]. Manual workflows rarely support the volume of emulations needed to estimate institution-level patterns reliably.

Recent large language model (LLM) systems have begun to automate components of trial emulation, raising the possibility of scalable, standardized replication [72,73,74,75]. Agent-based systems, even outside the RCT setting, are able to intake clinical text, phenotype ontology terms, and genetic data suggesting new depths for emulation and trial design [80]. Yet automation alone does not solve the interpretability problem: even if an agent can generate emulation results, the field lacks a principled way to treat the pattern of agreement and disagreement across emulations as a learnable object, rather than as isolated successes or failures. We argue that this pattern can be modeled explicitly as a decomposition: (i) a comparison-specific expectation of reproducibility informed by the published literature, and (ii) an institution-specific systematic shift capturing how local data-generating processes differ from published benchmarks, plus residual heterogeneity that bounds predictability.

In atrial fibrillation anticoagulation, this issue is clinically consequential because treatment choice depends on net benefit, specifically reductions in ischemic stroke and systemic embolism weighed against excess serious bleeding [31,32,33]. Multiple mechanisms can alter the realized effectiveness and safety of anticoagulants across health systems, including variability in warfarin management (e.g., time in therapeutic range) [43,44], off-label direct oral anticoagulant dosing, differential persistence [37,38], and outcome ascertainment differences [48,49]. Discrepancies between RCT and EHR estimates therefore directly affect decision-relevant quantities and are plausible manifestations of real system differences, not merely analytic shortcomings.

In this study, we present an agentic framework that enables cumulative institutional learning from trial emulation and published literature. The framework operationalizes the view that literature, EHRs, and RCTs comprise a measurement system through which a latent drug effect is expressed under different observational regimes. To obtain and synthesize these knowledge sources, we deploy Biomni, an open-source autonomous LLM agent, to execute a fully specified end-to-end EHR trial emulation pipeline within an OMOP-mapped database, including automated construction of literature-informed priors. These components are integrated under a Bayesian hierarchical calibration model that estimates both the institution-level transformation of trial evidence and the latent drug effects themselves. The result moves beyond emulation to a calibrated, uncertainty-aware estimate of how treatment effects are expected to manifest within the local care environment.

## Methods

### Study overview

We developed and evaluated an agentic framework for EHR-based emulation of randomized clinical trials (RCTs) and for learning structured patterns of discrepancy between emulated and published trial results. The framework comprises two stages: (1) autonomous, instruction-driven trial emulation using an LLM agent operating against an OMOP Common Data Model (CDM) EHR, and (2) Bayesian calibration that decomposes EHR–RCT discrepancies into a literature-informed, comparison-specific reproducibility expectation and an institution-specific systematic shift with residual heterogeneity. Performance was assessed using leave-one-out cross-validation across four in-domain trials and a separate out-of-distribution evaluation. Additional implementation details are provided in Supplementary Methods (Sections S1–S9).

### Data source and trial selection

EHR data were obtained from the Mount Sinai Health System and mapped to OMOP CDM version 5.4 using standardized vocabularies (RxNorm, SNOMED CT, LOINC) [46]. Data were accessed only within the institutional secure computing environment (Supplementary Methods S1). We selected five atrial fibrillation anticoagulation RCTs. The primary calibration set comprised four DOAC-versus-warfarin trials—ARISTOTLE [1], ROCKET AF [4], RE-LY [2], and ENGAGE AF–TIMI 48 [3]—chosen for a shared comparator (warfarin), a common primary endpoint (stroke or systemic embolism), and well-characterized published results. AVERROES [5] (apixaban versus aspirin) served as an out-of-distribution test due to different comparator and clinical context. Published hazard ratios and confidence intervals were extracted and converted to log–hazard ratios and standard errors (Supplementary Methods S2).

### Agentic trial emulation

We deployed Biomni, an autonomous LLM agent designed to execute multi-step biomedical workflows via instruction-following, tool use, and code generation/execution. The agent was run on the Minerva high-performance computing cluster with access to OMOP CDM tables stored in Apache Parquet format (Supplementary Methods S3). For each trial, Biomni executed a standardized end-to-end emulation pipeline: protocol parsing, phenotype and OMOP concept-set construction, cohort building, covariate extraction, confounder adjustment [20,21], treatment effect estimation, discrepancy computation, discrepancy diagnosis, and—when indicated—refinement and re-emulation. To quantify variability arising from agent stochasticity and analytic degrees of freedom, the full pipeline was executed three independent times per trial, each run starting from the same instruction documents and data but proceeding without access to other runs (Supplementary Methods S4–S5).

### Emulation outputs and discrepancy definition

For each run and trial, the agent produced (i) an EHR-derived treatment effect estimate on the log-hazard ratio scale, \hat{\tau}^{EHR}, with standard error SE^{EHR}; (ii) the corresponding published RCT log-hazard ratio, \tau^{trial}=\log(HR), with standard error derived from the reported confidence interval; and (iii) structured artifacts documenting concept sets, cohort sizes, event counts, covariates, model diagnostics, and failure modes (Supplementary Methods S5). We defined the run-level discrepancy as

\delta = \hat{\tau}^{EHR} - \tau^{trial}, \quad SE_\delta = \sqrt{(SE^{EHR})^2 + (SE^{trial})^2}.

Prespecified quality flags (e.g., sparse events, poor covariate balance, implausible discrepancies) were generated to support interpretation and sensitivity analyses (Supplementary Methods S5).

### Literature-informed priors for expected disagreement

For each drug comparison, the agent constructed a literature-informed expectation for EHR–RCT disagreement (reproducibility expectation). The agent conducted structured searches prioritizing direct evidence (published emulations/observational studies of the same comparison) and then indirect/meta-epidemiological evidence. Extracted discrepancy/bias estimates on the log-HR scale were pooled using random-effects meta-analysis [25,26] with an explicit domain-transfer uncertainty term to account for setting differences, yielding per-comparison \mu_{lit,k} and \sigma_{lit,k} (Supplementary Methods S6).

### Bayesian calibration model

We modeled each trial’s causal effect as a latent quantity \tau_k on the log-HR scale, observed through the published RCT channel and the local EHR emulation channel. For trial k,

\tau_k \sim \mathcal{N}(\mu_{lit,k}, \sigma^2_{lit,k}),

\hat{\tau}^{trial}_k \sim \mathcal{N}(\tau_k, (SE^{trial}_k)^2),

\hat{\tau}^{EHR}_{r,k} \sim \mathcal{N}(\tau_k + \mu_{site}, (SE^{EHR}_{r,k})^2 + \sigma^2),

where r indexes the run, \mu_{site} is an institution-level systematic shift shared across comparisons, and \sigma captures residual heterogeneity beyond reported standard errors. We used weakly informative priors \mu_{site}\sim \mathcal{N}(0,0.20) and \sigma\sim Half-Normal(0.20), and fit the model in PyMC using MCMC [27,30] (Supplementary Methods S7). The calibrated local effect for trial k is \tau^{local}_k=\tau_k+\mu_{site}.

### Evaluation and sensitivity analyses

We assessed calibration using leave-one-out cross-validation [58] across the four DOAC-versus-warfarin trials: in each fold, all three run-level observations for the held-out trial were removed, the model was fit on the remaining trials, and the posterior predictive distribution for the held-out trial was computed. We compared uncalibrated and calibrated mean absolute error (MAE) relative to published trial effects and computed empirical coverage of 95% posterior predictive intervals [28,29]. AVERROES was evaluated as out-of-distribution using the model fit on all four DOAC trials. Sensitivity analyses varied calibration priors and the domain-transfer uncertainty used in literature prior construction (Supplementary Methods S8).

### Human oversight and autonomy boundaries

Instruction documents were authored by the investigators prior to agent execution. During execution, Biomni operated without human review or correction between pipeline stages; investigators did not edit concept sets, adjust cohort logic, add clinical knowledge, or tune modeling parameters in response to intermediate results. Limited human involvement was restricted to operational facilitation needed to complete runs as specified in the written instructions (e.g., providing file path references required by the instructions, or re-invoking a step when the agent produced an empty/incomplete output due to runtime failure). After completion of all trials and runs, investigators reviewed generated artifacts for clinical plausibility and data integrity, and the Bayesian calibration analysis was specified and fit using the agent outputs as inputs. Expanded documentation of oversight actions, logged artifacts, and reproducibility materials is provided in Supplementary Methods S9.

### Ethics and governance

The study was approved by the Institutional Review Board of the Icahn School of Medicine at Mount Sinai (protocol number). All analyses were conducted within the HIPAA-compliant institutional environment, including the use of Biomni. Use of LLM-based tools is disclosed; instruction documents, agent artifacts, and analysis code are provided in the Supplementary Appendix.

## Results

### Execution of autonomous emulations and replicated runs

Biomni executed three independent end-to-end runs of the full emulation pipeline across the four in-domain DOAC-versus-warfarin trials, ARISTOTLE [1] (apixaban), ROCKET AF [4] (rivaroxaban), RE-LY [2] (dabigatran), and ENGAGE AF–TIMI 48 [3] (edoxaban), producing 12 run-level EHR effect estimates on the log–hazard ratio scale and corresponding EHR–RCT discrepancies. A fifth trial, AVERROES [5] (apixaban versus aspirin), was emulated separately and reserved for out-of-distribution evaluation. Because each run represents an independent realization of the emulation task under identical instructions, pooling across runs yielded 9 training observations per leave-one-out fold (3 runs × 3 trials), stabilizing calibration in the small-trial setting (Supplementary Table SR1; Supplementary Figure SR1).

### Literature-informed priors for expected EHR–RCT disagreement

For each drug comparison, the agent conducted structured literature retrieval and constructed comparison-specific priors encoding expected disagreement between observational and randomized evidence. The agent retrieved 174–560 records per comparison and extracted effect estimates from observational studies of DOACs versus warfarin in atrial fibrillation [34,35,36]. Across all four comparisons, the resulting priors favored greater apparent DOAC benefit in observational data than in the corresponding RCTs, with prior means \mu_{lit} ranging from −0.572 for apixaban to −0.391 for edoxaban (Table 1; Supplementary Table SR2). Prior dispersion differed across drugs: apixaban showed the widest spread (\sigma_{lit}=0.306), whereas rivaroxaban was more tightly constrained (\sigma_{lit}=0.134) (Table 1; Supplementary Figure SR2).

**Table 1:** Emulation Characteristics: Best Agent Run Per Trial.

| Trial | Run | $n_{\text{exp}}$ | $n_{\text{comp}}$ | Method | Covariates | Max SMD |
| --- | --- | --- | --- | --- | --- | --- |
| ARISTOTLE | 1 | 32,208 | 12,500 | IPTW Cox PH | Age, sex, prior anticoag | 0.006 |
| ENGAGE AF | 3 | 142 | 22,480 | IPTW Cox PH | Age, sex | 0.335 |
| RE-LY | 2 | 2,738 | 20,690 | Crude log-IRR<sup>†</sup> | — | — |
| ROCKET AF | 1 | 12,941 | 12,919 | IPTW Cox PH | Age, sex, prior anticoag | 0.007 |

<sup>†</sup> Crude log incidence rate ratio (person-time, 0.5 continuity correction). The log-IRR approximates the log-HR when event rates are low and censoring is non-differential. No agent run achieved propensity score adjustment for dabigatran.
Selected from autonomous agent runs 1–3 based on: IPTW availability > covariate balance > SMD < 0.05 > sample size.

### Calibration performance in pooled leave-one-out validation (in-domain)

In pooled leave-one-out cross-validation across the four in-domain trials, Bayesian calibration reduced mean absolute error (MAE) from 0.567 to 0.224 log–hazard ratio (60.5% reduction) (Figure 1; Supplementary Table SR3). All four held-out trial effects fell within the 95% posterior predictive interval (100% empirical coverage; 4/4), and in each fold the calibrated prediction moved in the correct direction toward the published RCT result. No sampling divergences occurred in any fold.

**Figure 1:**
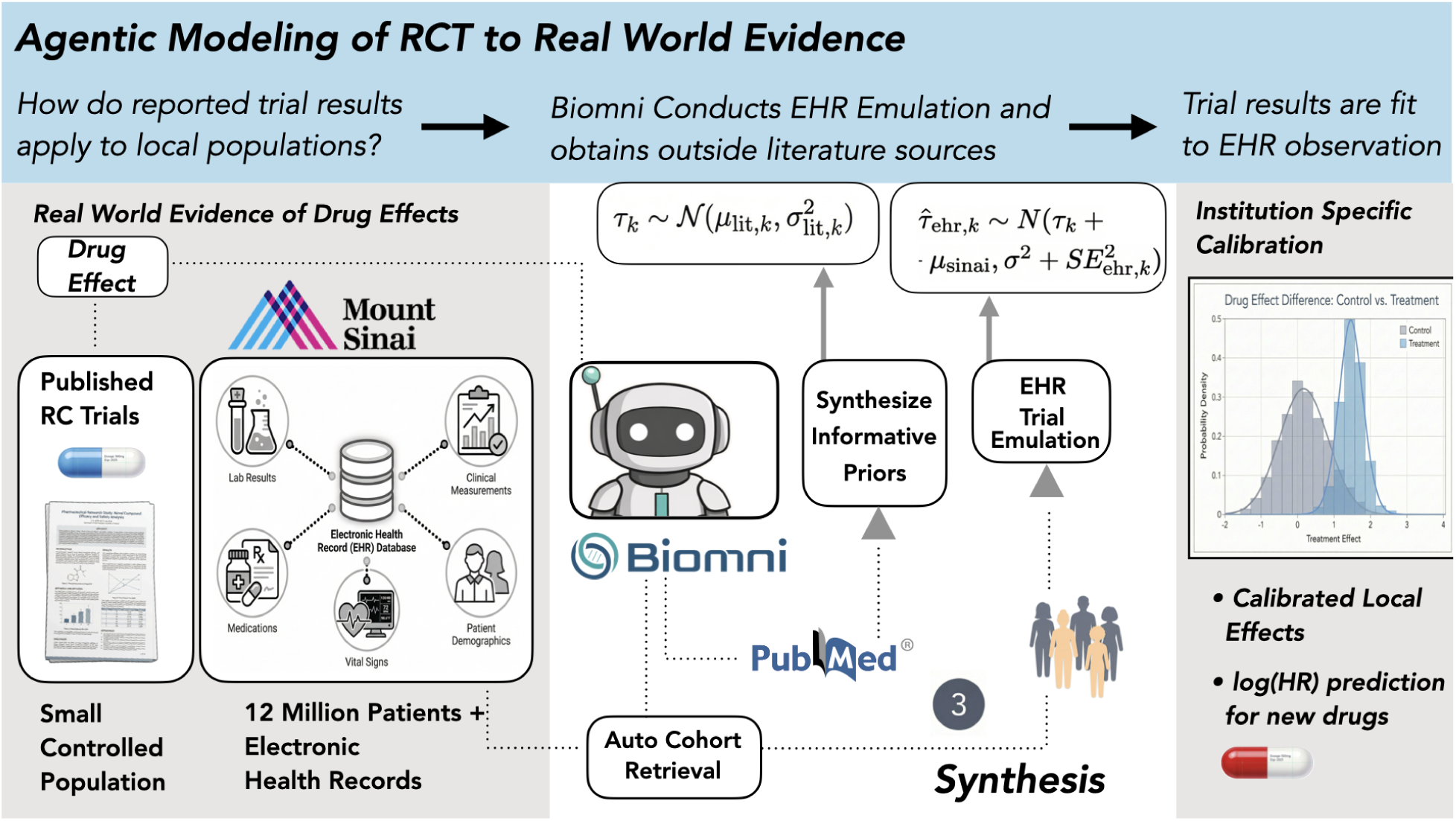
Drug effects reported in randomized clinical trials are not always reproduced by trial emulations using electronic health records. Biomni agent can receive clinical trial results and interface with private medical records databases. The agent autonomously reproduces the trial, extracting cohorts and assigning them to study arms and recording discrepancies. Additionally the agent retrieves related literature and synthesizes an informative prior of drug effects. Lastly, the trial results are integrated with the informative prior by a hierarchical bayesian model to learn local credible intervals for anticipated drug effects in the local health system through calibration.

Per-trial results illustrate the model’s behavior across discrepancy magnitudes (Table 2; Supplementary Figure SR4). ENGAGE AF exhibited the largest pooled uncalibrated error (1.247 log–HR), driven by a run-level outlier estimate; calibration reduced this error to 0.016, yielding a near-exact recovery of the published trial effect. ARISTOTLE showed the largest consistent discrepancy across runs; calibrated error was 0.212, with the published effect comfortably contained within the 95% credible interval (−0.625 to 0.259). RE-LY was the most challenging case: it had the smallest uncalibrated discrepancy (0.187), and calibration over-corrected (calibrated error 0.316), although the published effect remained within the 95% credible interval (−0.787 to −0.102).

**Table 2:** Informative Literature Priors Used in Calibration.

| Trial | Drug | $\mu_{\text{lit}}$ | $\sigma_{\text{lit}}$ | Direction | Interpretation |
| --- | --- | --- | --- | --- | --- |
| ARISTOTLE | Apixaban | -0.572 | 0.306 | RWD > benefit | Strong obs. overestimate |
| ENGAGE AF | Edoxaban | -0.391 | 0.144 | RWD > benefit | Moderate obs. overestimate |
| RE-LY | Dabigatran | -0.458 | 0.154 | RWD > benefit | Moderate obs. overestimate |
| ROCKET AF | Rivaroxaban | -0.404 | 0.134 | RWD > benefit | Moderate obs. overestimate |

$\mu_{\text{lit}}$ : expected log-HR discrepancy ( $\tau_{\text{ehr}} - \tau_{\text{trial}}$ ) from published observational studies of each drug vs warfarin in AF. Negative values indicate the literature expects RWD to show greater DOAC benefit than the RCT.
$\sigma_{\text{lit}}$ : dispersion of extracted observational effect estimates across sources (log-HR scale). Used as the literature prior SD in the calibration model.

**Table 3:** Agent Selected Literature for Constructing Informative Prior.

| Trial | Drug | PMIDs<br>Retr. | Drug-Uniq. | Top-100<br>Cov. | Example Drug-Unique PMIDs |
| --- | --- | --- | --- | --- | --- |
| ARISTOTLE | Apixaban | 482 | 106 | 97% | 41554390 41240461 40846025 |
| ENGAGE AF | Edoxaban | 174 | 33 | 69% | 40483337 35976428 36513051 |
| RE-LY | Dabigatran | 559 | 121 | 99% | 37306492 35294623 36284318 |
| ROCKET AF | Rivaroxaban | 560 | 90 | 100% | 41632318 39585535 38496142 |

### Posterior estimates of institutional shift and residual heterogeneity

Across leave-one-out folds, the posterior for the institution-specific shift \mu_{site} was consistently positive, with medians ranging from 0.364 (ARISTOTLE held out) to 0.580 (ROCKET AF held out) and a mean posterior standard deviation of 0.103 (Figure 2; Supplementary Table SR6). This indicates systematic attenuation of estimated DOAC benefit in the institutional EHR relative to published RCT results beyond what the comparison-specific literature priors predicted. Residual heterogeneity \sigma had posterior medians ranging from 0.199 to 0.264 across folds (mean 0.240; mean posterior SD 0.064), reflecting moderate trial-to-trial variation unexplained by the literature priors, the shared institutional shift, or reported standard errors (Supplementary Table SR6; Supplementary Figure SR6).

**Figure 2:**
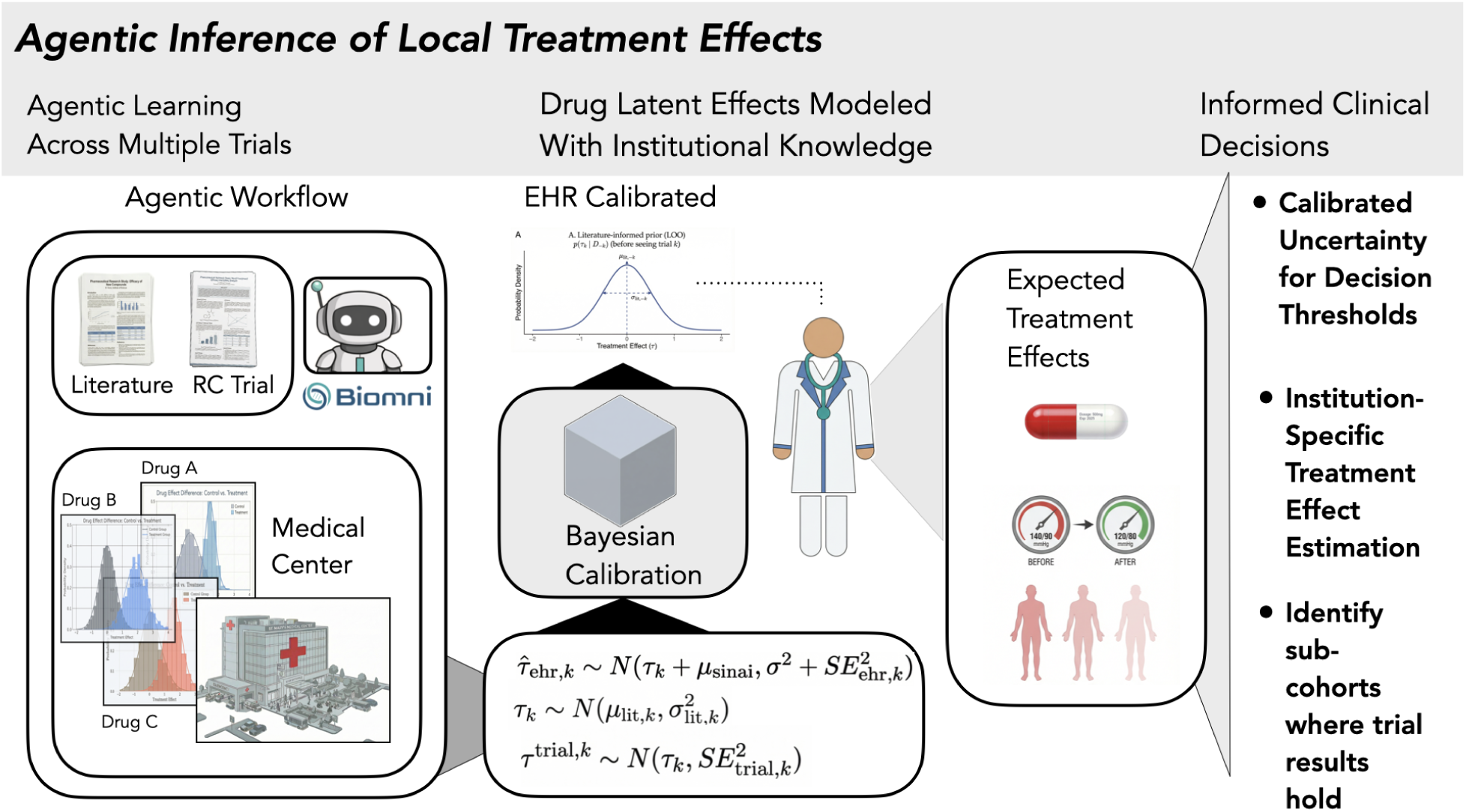
When the agent is applied to multiple different clinical trials for drugs in related or potentially unrelated clinical classes, divergence of observed treatment effects from published literature and RCT results allow a model to learn anticipated institutional factors that produce valid local credible intervals for drug effects. Clinicians can use these intervals to ground RCT results with the clinical reality of local patients allowing for more effective net benefit estimations and for identifying both individuals likely to respond similarly to a trial and for whom there is less likelihood. This allows for automated learning of whole health system treatment effects.

### Out-of-distribution evaluation

We next tested whether the learned transport structure generalized beyond the DOAC-versus-warfarin domain. Trained on all pooled in-domain observations, the model was applied to AVERROES [5] (apixaban versus aspirin) without retraining. Uncalibrated prediction error was 0.379 log–HR; after calibration, error decreased to 0.051 (86.5% reduction), and the published trial result (log–HR = −0.799) fell within the 95% credible interval (−1.487 to −0.197) (Figure 3; Supplementary Table SR7). When the model was trained using Run 2 alone, calibrated error was 0.138 and the published effect was also covered, consistent with improved stability under pooled training.

**Figure 3:**
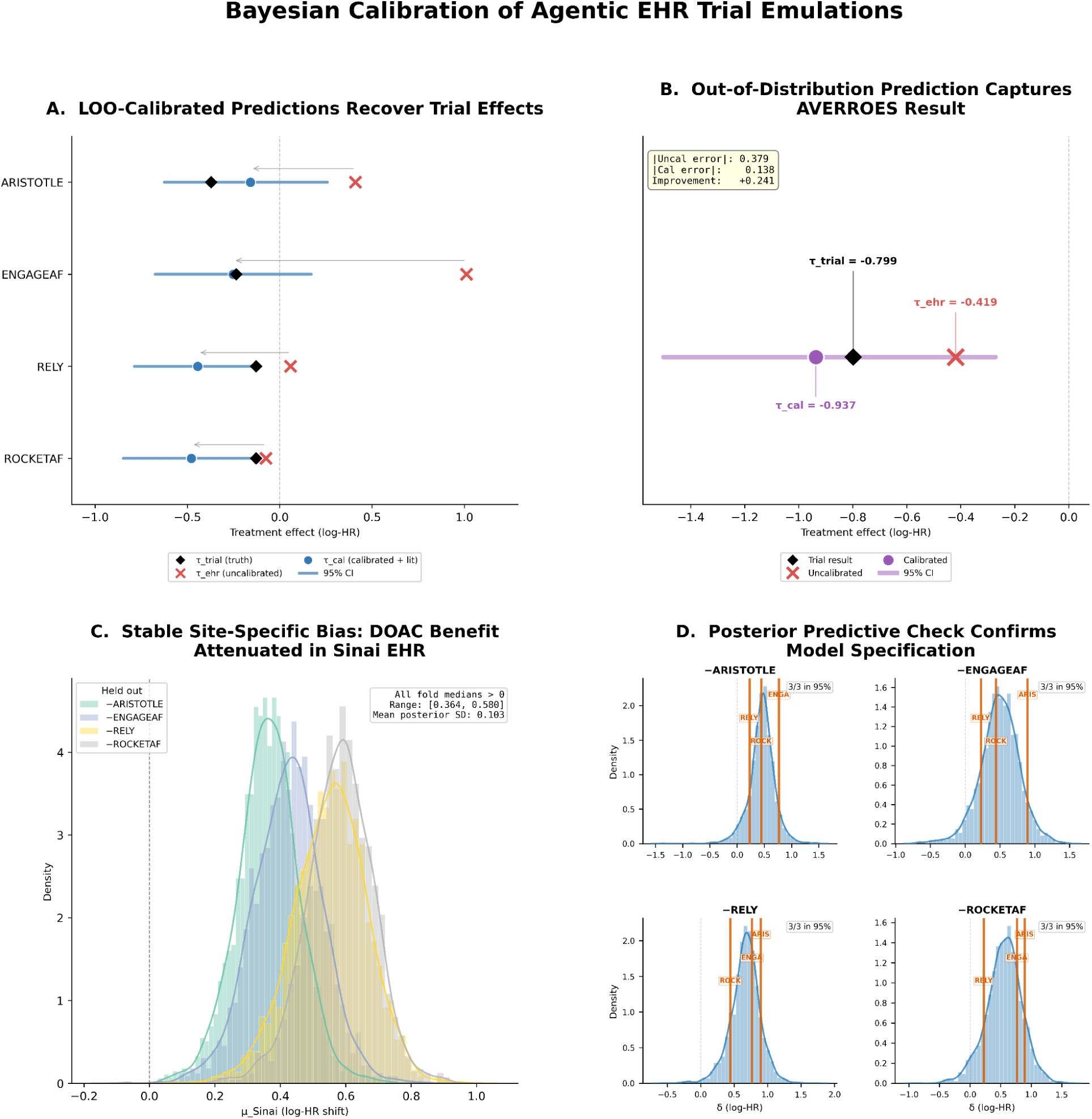
A. We show that integrating agent-derived priors with EHR emulations across trials produces calibrated 95% credible intervals (blue) that adjust away from the raw EHR estimate (red X) and generally contain the randomized trial result (black diamond), with interval width reflecting uncertainty about transportability and the range of effects plausibly realized in the local health system. The predicted (held out) trial result is indicated in the Y-axis (from training on 3 included trials). Because the raw EHR emulation does not account for systematic institutional bias or excess heterogeneity, it cannot be interpreted as the expected local causal effect; the calibrated posterior interval instead defines the plausible range of that effect after integrating all sources of uncertainty. B. Trial results from unobserved comparators (here Aspirin) from AVERROES are also successfully predicted using the institutional prior learned from previous training. C. Posterior distributions of the institution-specific bias parameter (μ_Sinai) estimated in leave-one-out analyses. Across held-out trials, the bias parameter is consistently positive, indicating systematic attenuation of DOAC benefit in the Sinai EHR relative to randomized trial effects. The stability of these posteriors across trials supports the assumption of a shared site-level calibration parameter. D. Posterior predictive distributions for trial-specific discrepancies under the calibrated model. Observed trial–EHR differences lie within the 95% predictive intervals for all held-out trials, indicating that the hierarchical bias and variance structure adequately captures cross-trial heterogeneity.

### Agent degrees of freedom and run-to-run variability

The agent exercised analytic judgment at 26 documented decision points spanning exposure definition, outcome concept selection, covariate selection, modeling strategy, and quality assessment. These choices varied across runs; for example, the agent selected different covariate sets for propensity score estimation in ROCKET AF and invoked different modeling fallbacks when preferred specifications failed convergence. Treating run-level outputs as exchangeable measurements and pooling across runs improved calibration performance relative to per-run training, consistent with the use of replicated executions to quantify and partially average over agent-induced analytic variability (Supplementary Table SR8; Supplementary Figure SR5).

## Discussion

For a clinician choosing between apixaban and warfarin, the relevant quantity is not simply the ARISTOTLE hazard ratio [1], but how that effect plays out within the institution’s patients, prescribing practices, and outcome measurement regime. Disagreement between trial and EHR estimates of anticoagulation efficacy is structured, learnable, and useful for decision-making. Across four DOAC trials, the model identified a stable institution-level shift and generalized to a structurally distinct out-of-distribution comparison. Although we do not estimate patient-specific effects, the calibrated posterior yields institution-appropriate uncertainty that can inform individual decisions insofar as those decisions depend on institution-level influences.

These findings challenge the prevailing paradigm in trial emulation, which treats divergence between EHR and RCT estimates as analytic failure to be minimized. TRIALSCOPE pushed against this, combining language models, probabilistic harmonization, and causal survival modeling to emulate trials at scale [76], reproducing nine of eleven lung-cancer hazard ratios with extensive diagnostic validation — impressive work in single-emulation fidelity.

Yet concordance and discrepancy remain under-interpreted. For example, TRIALSCOPE’s emulation of FLAURA [76] yields a hazard ratio of 0.61 versus a published 0.63, which is near-perfect agreement. But the Providence and trial populations differ in mutation subtypes, prior therapy, and comorbidity burden, raising the question of whether agreement reflects genuine transportability [15,16,19] or compensating biases [7]. Diagnostic suites primarily assess internal model behavior rather than external validity.

Our point is not that discrepancy necessarily implies bias; it is that without a joint model of system-level expression, discrepancy and concordance are both under-interpreted. Passing internal diagnostics does not explain why agreement occurs or what divergence signifies. This reflects a structural gap. Large initiatives such as RCT DUPLICATE [42] report concordance across trials, but emulations are typically evaluated independently. The pattern of discrepancies across trials, including non-trial literature sources, is rarely modeled jointly. The unasked question is: what does the structure of divergence reveal about the observation system itself?

Rather than minimizing discrepancies in isolation, we treat their pattern across trials as data. The literature prior μ_lit,k encodes expected reproducibility for each comparison; the site-specific shift μ_Sinai captures systematic deviation from that expectation. Instead of asking whether an emulation achieved equivalence, we estimate how, and in what direction, the local system reshapes external evidence, with propagated uncertainty. One emulation is a case report; multiple emulations analyzed jointly characterize how clinical evidence behaves within a care environment.

The model posits that neither the trial nor the EHR estimate is ground truth. Both are noisy projections of a latent treatment effect, τ_k, defined independently of any measurement regime [62,63,64]. The RCT observes this effect under enrollment and adjudication constraints; the EHR observes it after it passes through institutional selection, prescribing, and outcome capture processes. Divergence is therefore two system-conditioned measurements of the same causal quantity, not truth versus approximation.

Each discrepancy is decomposed into three components: a literature-informed expectation μ_lit,k, an institution-level shift μ_Sinai shared across comparisons, and residual heterogeneity. This separates drug-level reproducibility from system-level transformation. Only divergence beyond literature expectation informs μ_Sinai — a key distinction in small-N calibration.

Although the statistical structure resembles hierarchical partial pooling, what is pooled is not effects toward a grand mean but discrepancies toward an explicit model of how evidence is expressed locally. The novelty lies in treating divergence as structured information about the observation system.

For each drug comparison, the literature prior encodes the empirical distribution of published EHR–trial discrepancies. Across the four DOAC trials, the agent-constructed priors favored greater apparent benefit in observational data than in RCTs (μ_lit from −0.572 for apixaban to −0.391 for edoxaban), consistent with many real-world analyses reporting more favorable hazard ratios for DOACs versus warfarin [34,35,36]. Dispersion varied: apixaban showed substantial heterogeneity (σ_lit = 0.306), whereas rivaroxaban was more tightly constrained (σ_lit = 0.134).

The prior serves as an inferential baseline, distinguishing broadly observed reproducibility patterns from institution-specific effects. Because μ_lit,k is derived from published discrepancies, it may reflect reporting and publication biases [9,14]; incorporating registries and negative findings would strengthen this layer.

After accounting for literature expectations, the model identified a persistent institution-level shift. The posterior for μ_Sinai was consistently positive across leave-one-out folds, with medians ranging from 0.364 to 0.580 and mean posterior standard deviation of 0.103, indicating systematic attenuation of DOAC benefit relative to trial estimates. Stability under perturbation supports interpretation as a genuine system property.

Such attenuation plausibly reflects high-quality warfarin management [43,44], dose modification, differential persistence [37,38], comorbidity burden [40,41], or outcome ascertainment differences [48,49]. These are not confounders to eliminate [23,24] but features of the care environment through which biological effects are expressed [15,16,19]. The institutional shift quantifies their aggregate consequence.

Residual heterogeneity remained after accounting for drug-level and institutional components, capturing variation not explained by stable system effects. Modeling this component prevents overconfidence and ensures calibrated estimates reflect both accumulated knowledge and its limits.

System-level effects cannot be estimated from a single emulation. The site-level shift becomes identifiable only through accumulation of standardized discrepancies across trials. Literature-informed priors likewise require systematic retrieval and harmonization.

The agentic workflow enables this accumulation. Analytic judgment remains present but is logged and reproducible. Between-run variability becomes observable rather than hidden; pooling across runs converts analytic flexibility into estimable uncertainty. Automation thus enables institutional learning rather than merely accelerating individual emulations.

A harder test is whether learned system properties extend beyond training comparisons. Calibrated exclusively on DOAC-versus-warfarin trials, the model reduced prediction error when applied to AVERROES [5] (apixaban versus aspirin) and produced uncertainty intervals containing the published effect. A single out-of-distribution comparison is not definitive, but it supports the interpretation that the model learned a reusable system-level transformation rather than comparator-specific adjustments.

The model assumes additive decomposition and temporal stability of institutional effects. Yet more complex interactions or drift may exist. With four training trials, the model operates near its identifiability floor. Stability of μ_Sinai under leave-one-out perturbation mitigates overfitting concerns, but we view this as a demonstration of feasibility. Parameters should stabilize as additional comparisons accrue. We calibrate efficacy only; extending to bleeding and composite net-benefit endpoints is necessary for decision-relevant translation. Incorporating physiological pathways or mechanisms of action into the hierarchy would allow partial pooling across biologically related drugs, improving efficiency and interpretability as the framework scales.

Beyond these extensions, the framework’s emphasis on system-level learning also distinguishes it from an alternative paradigm for individualized evidence generation.Digital twin frameworks estimate individualized counterfactual outcomes by constructing patient-specific replicas under alternative treatment assignments. While powerful for modeling biological heterogeneity [78], they do not provide a principled mechanism for learning how published randomized evidence is transformed as it enters routine clinical care. Nor do they represent institution-level forces such as prescribing culture, adherence, and outcome ascertainment that operate above the individual yet materially shape observed treatment effects.

The present framework addresses a different inferential problem. We do not directly estimate individualized treatment effects. We generate a system-level estimate to inform decision-making and treat this transformation as a learnable statistical object. By estimating an institution-specific transport parameter shared across drug comparisons, we characterize how the local data-generating and measurement regime shifts treatment effects relative to randomized benchmarks and thereby construct a prior for local inference. For example, signaling when anticoagulant trial results do not directly transport to the institution. Such signals prompt examination of the institutional factors underlying divergence and the patient specific evidentiary basis for prescribing in that context.

More fundamentally, we hold that treatment effects are not properties of patients alone but of patients embedded within care regimes. External validity and transportability theory formalize that causal effects are regime-indexed[77]: when the regime changes, the effect may change. Patient-specific variables alone are therefore insufficient to characterize expected benefit. Institutional context must be considered alongside biological heterogeneity when interpreting treatment effects. In this view, institutional context is not a confounder to eliminate but part of the inferential prior through which evidence is interpreted and clinical decisions are made.

Future extensions include mechanism-informed hierarchies, cross-system learning, and subgroup-level calibration [79] - the latter representing a natural convergence point at which system-level transport and individual heterogeneity can be jointly modeled. The central insight remains: divergence is not something to eliminate but something to model, and it tells us how treatments operate across populations and care environments.

## CONCLUSIONS

A published trial result enters clinical practice as an implicit prior about treatment effect. Clinicians often recognize that their patients do not respond exactly as trials predict, yet lack a formal mechanism to reconcile those observations with the evidence guiding care. This study provides that mechanism. The calibrated posterior yields not a single adjusted number but a distribution: a locally grounded estimate of effect, the uncertainty introduced by system-level expression, and the probability that benefit meaningfully persists. From this distribution, a clinician can assess whether expected benefit clears the threshold that justifies treatment in their population and how confident that judgment should be.

Understanding how evidence plays out in practice cannot be achieved through a single emulation. It requires repeated, standardized replication, systematic synthesis of prior evidence, and pooled analysis across comparisons, a scale that is not practical without automation. The agent enables this accumulation. As discrepancies accrue, their pattern becomes informative: which trial results reproduce locally and which attenuate reveals how patient mix, prescribing practices, and outcome capture shape observed effects. With sufficient scale, this learning progresses beyond a single global adjustment toward identifying structured sources of divergence and subgroups in which trial expectations are more or less likely to hold. In doing so, it transforms clinical intuition about “what works here” into a measurable property of the health system, establishing a feedback loop in which clinicians and autonomous agents jointly refine expectations about treatment effect as evidence evolves.

## Data Availability

The electronic health record data used in this study contain protected health information and cannot be shared publicly under HIPAA regulations and institutional policy. Access to deidentified data may be available upon reasonable request to the corresponding author, subject to approval by the Institutional Review Board of the Icahn School of Medicine at Mount Sinai and execution of a data use agreement. Published RCT results used for calibration are publicly available: ARISTOTLE (PMID: 21870978), RELY (PMID: 19717844), ENGAGE AFTIMI 48 (PMID: 24243014), ROCKET AF (PMID: 21830957), and AVERROES (PMID: 21090905).

## Supplementary Materials

### Supplementary Methods

#### S1. Data source and OMOP CDM implementation

EHR data were obtained from the Mount Sinai Health System and mapped to the Observational Medical Outcomes Partnership (OMOP) Common Data Model (CDM) version 5.4 [46]. The CDM uses standardized vocabularies (RxNorm, SNOMED CT, LOINC) to represent drug exposures, conditions, procedures, and measurements across sites and time. OMOP vocabulary files and clinical data tables were stored as Apache Parquet files within the institutional secure computing environment on the Minerva high-performance computing cluster.

The agent queried both vocabulary and clinical tables. Vocabulary tables included concept, concept_ancestor, and concept_relationship. Clinical tables included person, drug_exposure, condition_occurrence, measurement, procedure_occurrence, observation_period, and death (where available). For each emulation run, all executed queries, tables accessed, and intermediate outputs were written to run-specific artifacts.

#### S2. Trial selection and extraction of published trial results

We selected five randomized trials in atrial fibrillation anticoagulation.

**Calibration set (in-domain):** four landmark direct oral anticoagulant (DOAC) trials with a shared comparator (warfarin) and common primary efficacy endpoint (stroke or systemic embolism):

- ARISTOTLE [1] (apixaban vs warfarin)
- ROCKET AF [4] (rivaroxaban vs warfarin)
- RE-LY [2] (dabigatran vs warfarin)
- ENGAGE AF–TIMI 48 [3] (edoxaban vs warfarin)

These trials were chosen because they share key structural elements—comparator, endpoint family, and well-characterized published results—making them suitable for leave-one-out evaluation of a calibration model with limited trial count.

### Out-of-distribution test

AVERROES [5] (apixaban vs aspirin) was included to evaluate generalization outside the DOAC-versus-warfarin domain. AVERROES differs from the calibration set in comparator (aspirin rather than warfarin) and enrollment context (patients deemed unsuitable for warfarin), providing a stringent test of transport beyond training comparisons.

### Extraction and transformation

Published hazard ratios (HRs) and 95% confidence intervals were extracted from trial manuscripts. Effect sizes were converted to the log-hazard ratio scale as:

\tau^{trial} = \log(HR),

\quad

SE^{trial} = \frac{\log(HR_{upper}) - \log(HR_{lower})}{3.92}.

Where event counts, follow-up windows, or key eligibility details were available, these were recorded as part of the trial extraction artifact used by the agent in downstream steps.

## S3. Biomni agent architecture and deployment

Biomni is an open-source LLM agent built on LangChain and LangGraph, designed to execute multi-step biomedical workflows through instruction-following, tool use, and code generation/execution. The agent operates by (i) receiving a structured instruction document specifying inputs, procedures, and required outputs; (ii) decomposing tasks into executable steps; (iii) generating and running Python code against target data resources; and (iv) producing structured outputs (CSV/YAML) conforming to specified schemas. Biomni maintains a state graph to track progress across tasks and includes error handling and fallback strategies. For this study, the agent was deployed on the Minerva high-performance computing cluster with access to a subset of the institutional OMOP CDM v5.4 database stored in Apache Parquet format. The agent was backed by GPT-5.2 (OpenAI; accessed via Azure OpenAI API, May–June 2025) and communicated with the language model via Azure OpenAI. No component of the agent was trained or fine-tuned on the trials under study; it operated using general-purpose language modeling capabilities and the structured instruction documents described below.

## S4. Instruction-driven emulation pipeline and replicated runs

### S4.1 Instruction documents and execution model

The agent was guided by six sequential instruction documents, each specifying a discrete phase of the emulation pipeline. The instruction documents were authored by the study investigators prior to execution and provided to the agent as markdown files. Each instruction specified (i) required inputs (file paths, schemas, prior artifacts), (ii) analytical procedures, (iii) fallback strategies in the event of failure, and (iv) exact output formats (YAML or CSV with defined schemas). The agent executed each instruction autonomously; outputs from each stage served as inputs to subsequent stages.

**Table S1. Summary of instruction-driven pipeline**

1\. Concept extraction and phenotype definition

2\. Cohort construction and emulation

3\. Literature-informed prior construction

4\. Discrepancy computation and quality flagging

5\. Discrepancy diagnosis

6\. Refinement and re-emulation

### S4.2 Replicated runs to quantify agent variability

To characterize variability introduced by the agent’s stochastic decision-making and analytic degrees of freedom, the full six-instruction pipeline was executed three independent times per trial (Runs 1–3). Each run began from the same instruction documents and the same underlying database but proceeded without access to other runs. Runs independently selected concept sets, constructed cohorts, selected covariates for propensity score models, and made modeling decisions under the same method hierarchy. The replicated runs were treated as exchangeable measurements of discrepancy for calibration and validation (Sections S7–S8). Per-run analyses were conducted to assess robustness to individual-run idiosyncrasies.

## S5. Detailed description of instruction stages

### S5.1 Instruction 1: Concept extraction and phenotype definition

For each trial, the agent read the published manuscript (PDF provided) and extracted core trial elements: intervention and comparator drugs, primary endpoint definition, eligibility criteria, and exclusion criteria. The agent then queried OMOP vocabulary tables (concept, concept_ancestor, concept_relationship) to translate each element into concept sets.

- **Drug exposures:** mapped at the RxNorm ingredient level, including all descendant concepts (formulations, strengths, branded products) via concept\_ancestor.
- **Conditions/diagnoses:** mapped to SNOMED CT concepts with descendant expansion.
- **Endpoint mapping:** operationalized using OMOP concepts representing trial endpoint components (e.g., stroke/systemic embolism), with documentation of ambiguity where trial endpoint definitions did not map cleanly to OMOP constructs.

For each concept set, the agent documented search terms used, candidate concepts considered, concepts selected, and any ambiguities or trade-offs. Published trial results were extracted and converted to the log-HR scale (Section S2).

### S5.2 Instruction 2: Cohort construction and emulation

Using concept sets from Instruction 1, the agent identified exposure and comparator cohorts from the OMOP drug_exposure table. The index date was defined as the first qualifying drug exposure.

#### Eligibility and exclusion

Trial eligibility and exclusion criteria were applied where operationalizable in EHR data; criteria not expressible in OMOP concepts or unavailable in structured EHR (e.g., investigator-assessed life expectancy) were documented as coverage gaps.

#### Follow-up

Follow-up began at index and continued until the earliest of: outcome event, death, end of observation period, drug discontinuation, or administrative censoring, consistent with the instruction document.

#### Baseline covariates

Covariates were extracted from records on or before index, including:

- demographics: age, sex
- comorbidities: history of stroke/TIA, major bleeding, heart failure, hypertension, diabetes, renal disease
- medications: prior anticoagulant use, antiplatelet use, proton-pump inhibitor use, statin use

Covariates were drawn from person, condition_occurrence, drug_exposure, and measurement tables.

#### Treatment effect estimation

The agent followed a prespecified method hierarchy:

1\. **IPTW (preferred):** propensity scores [20,21] via logistic regression of treatment assignment on baseline covariates; stabilized weights computed and truncated at 1st and 99th percentiles; weighted Cox proportional hazards model with treatment as the sole covariate; robust standard errors.

2\. **Covariate-adjusted Cox regression** 3\. **Unadjusted Cox regression**

4\. **Log-rank approximation**

#### Diagnostics and fallbacks

Covariate balance was assessed using standardized mean differences (SMDs) before and after weighting [22]. If the preferred method failed (e.g., non-convergence, singular matrix, missing library), the agent invoked the fallback sequence and documented the reason and method used.

Outputs included cohort sizes, events, follow-up summaries, model diagnostics, and \hat{\tau}^{EHR} with SE^{EHR}.

### S5.3 Instruction 3: Literature-informed prior construction

For each trial/comparison, the agent conducted a structured literature search to construct a prior for expected EHR–RCT discrepancy specific to that drug comparison. The search prioritized:

1\. **Direct evidence:** published emulations of the same RCT and/or observational studies of the same drug comparison and endpoint.

2\. **Indirect evidence:** RCT–observational concordance initiatives (e.g., RCT-DUPLICATE [42]) and meta-epidemiological evidence (e.g., Cochrane-style reviews quantifying RCT–observational differences [8,10]).

3\. **Methodological/domain evidence:** studies describing biases specific to anticoagulation observational research.

From each included source, the agent extracted or derived a point estimate of discrepancy/bias on the log-HR scale and its standard error. When studies reported HRs but not discrepancies, the agent derived discrepancies by subtracting the trial log-HR from the observational log-HR for a comparable endpoint where feasible, documenting assumptions.

Extracted estimates were pooled using a random-effects meta-analytic framework (DerSimonian–Laird [26]) with an additional domain transfer uncertainty term \tau_{domain} to account for differences between the literature setting and the target EHR environment (Section S6). The output was a per-comparison literature prior (\mu_{lit,k}, \sigma_{lit,k}) representing expected EHR–RCT discrepancy for each comparison.

### S5.4 Instruction 4: Discrepancy computation and quality flags

For each trial and run, the agent computed:

\delta_{r,k} = \hat{\tau}^{EHR}_{r,k} - \tau^{trial}_k,

\quad

SE_{\delta,r,k} = \sqrt{(SE^{EHR}_{r,k})^2 + (SE^{trial}_k)^2}.

The agent assigned prespecified quality flags based on thresholds including:

- SE^{EHR} \> 1.0 (uninformative estimate)
- fewer than 5 events in either arm (sparse events)
- |\\delta| \> 0.7 (implausible discrepancy)
- maximum post-weighting SMD \> 0.10 (poor balance)

Flag definitions and per-run flag outputs were recorded to support interpretation and sensitivity analyses.

### S5.5 Instruction 5: Discrepancy diagnosis

The agent generated a structured diagnostic assessment that reviewed:

- **Cohort validity:** arm sizes, event counts, index logic sanity checks, and cohort identity verification.
- **Modeling quality:** method used, balance, overlap, residual confounding risk, and reasons for fallback.
- **Concept mapping quality:** specificity/sensitivity tradeoffs in drug and outcome concept sets; eligibility coverage gaps.

Each discrepancy was evaluated against candidate explanations:

1\. residual confounding

2\. small-sample noise

3\. concept mapping error

4\. population mismatch

5\. duplicate or invalid cohort construction

6\. genuine EHR–RCT difference

For each, the agent recorded supporting evidence and an overall likelihood judgment.

### S5.6 Instruction 6: Refinement and re-emulation

Where the diagnostic report identified actionable refinements—such as revising concept sets, adding omitted covariates, or re-attempting a preferred modeling method—the agent implemented those changes, rebuilt cohorts, re-ran estimation, and recomputed discrepancies. Both original and refined estimates were preserved, along with a structured record of what was changed and why.

## S6. Literature prior pooling model and domain transfer uncertainty

Let b_i denote an extracted literature-based discrepancy estimate on the log-HR scale with standard error s_i. For each comparison k, estimates were pooled using a random-effects model with between-study variance \tau^2_{RE,k} estimated by the DerSimonian–Laird procedure [26]. To reflect uncertainty when transferring literature-derived discrepancy distributions to the local EHR environment, we added a domain-transfer variance component \tau^2_{domain}, yielding total dispersion:

\sigma^2_{lit,k} = \tau^2_{RE,k} + \tau^2_{domain} + \overline{s^2},

where \overline{s^2} denotes an average within-study variance contribution (as implemented in the agent’s pooling code). The resulting per-comparison prior mean \mu_{lit,k} and dispersion \sigma_{lit,k} were provided to the calibration model (Section S7). Sensitivity analyses varied \tau_{domain} over a prespecified range (Section S8).

## S7. Bayesian calibration model: specification, inference, and prediction

### S7.1 Model specification

For each trial k, we posit a latent treatment effect \tau_k representing the causal effect on the log-HR scale. The published trial result and the EHR emulation estimate are modeled as noisy observations of this latent effect through distinct channels.

#### Latent effect prior (literature-informed)

\tau_k \sim \mathcal{N}(\mu_{lit,k}, \sigma^2_{lit,k}).

#### RCT observation channel

\hat{\tau}^{trial}_k \sim \mathcal{N}(\tau_k, (SE^{trial}_k)^2).

#### EHR observation channel

\hat{\tau}^{EHR}_{r,k} \sim \mathcal{N}(\tau_k + \mu_{site}, (SE^{EHR}_{r,k})^2 + \sigma^2), where r indexes runs, \mu_{site} captures a persistent institution-level shift shared across comparisons, and \sigma captures residual trial-to-trial heterogeneity beyond reported standard errors.

The institution-expressed (local) effect is \tau^{local}_k := \tau_k + \mu_{site}.

### S7.2 Priors

We placed weakly informative priors on shared calibration parameters:

\mu_{site} \sim \mathcal{N}(0,0.20),

\quad

\sigma \sim \text{Half-Normal}(0.20).

Alternative prior specifications were examined in sensitivity analyses (Section S8).

### S7.3 Computation and diagnostics

The Bayesian calibration model was fit in PyMC (version 5) [30] using Markov chain Monte Carlo with four chains, 2000 iterations per chain (1000 warmup). Convergence was assessed via \hat{R} < 1.01 and effective sample size > 400 for all monitored parameters. Posterior predictive checks were conducted for each fit (summarized in output artifacts).

### S7.4 Prediction for held-out trials

For leave-one-out validation, holding out trial j removed all run-level EHR observations associated with that trial. The calibrated prediction for the held-out trial was the posterior predictive distribution of \tau_j (and \tau^{local}_j when reporting local effects), integrating uncertainty in \mu_{site}, \sigma, and measurement error. Point predictions were summarized using posterior means/medians as specified in evaluation scripts.

### S7.5 Identifiability note

Under K \ge 2 comparisons, the model parameters \mu_{site}, \sigma, and \{\tau_k\} are identifiable given (i) the anchoring effect of trial observations, (ii) informative comparison-specific literature priors, and (iii) variation in measurement error across EHR estimates. A formal identifiability argument and simulation-based sanity checks are provided in the Appendix.

## S8. Validation design, pooling strategy, and sensitivity analyses

### S8.1 Pooling across runs

Each run × trial combination contributed one discrepancy observation to the hierarchical calibration model. Pooling treats run-level results as exchangeable measurements arising from the same emulation task executed under identical instructions but with stochastic agent decisions. In leave-one-out cross-validation, holding out a trial removed all three of its run-level observations, and training proceeded on the remaining 9 observations (3 runs × 3 trials). Per-run analyses were also conducted to assess robustness to any single run.

### S8.2 Leave-one-out cross-validation (in-domain)

Calibration performance was assessed by leave-one-out cross-validation over the four DOAC-versus-warfarin trials. For each fold k = 1,\dots,4, the model was fit to the other three trials and used to predict the held-out trial. We compared:

MAE_{uncal} = \frac{1}{K}\sum_{k=1}^{K}\left|\tau^{trial}_k - \hat{\tau}^{EHR}_k\right|, \quad

MAE_{cal} = \frac{1}{K}\sum_{k=1}^{K}\left|\tau^{trial}_k - \hat{\tau}^{pred}_k\right|. Empirical coverage of 95% posterior predictive intervals was computed as the fraction of held-out trial effects falling within the interval. With K=4, coverage estimates are descriptive.

### S8.3 Out-of-distribution evaluation

For out-of-distribution evaluation, the model was fit to all four in-domain trials (12 run-level observations) and applied to AVERROES without retraining. Prediction error and coverage were computed analogously.

### S8.4 Sensitivity analyses

We assessed sensitivity of the calibration model to calibration priors by refitting under four specifications:

- **Base case:** \\mu\_{site} \\sim \\mathcal{N}(0,0.20), \\sigma \\sim Half-Normal(0.20)
- **Diffuse:** \\mathcal{N}(0,0.30), Half-Normal(0.40)
- **Skeptical:** \\mathcal{N}(0,0.05), Half-Normal(0.15)
- **Literature-heavy:** \\mu\_{site}\\sim \\mathcal{N}(0.06,0.06), \\sigma\\sim Truncated-Normal(0.18,0.06)

We also varied the domain transfer uncertainty \tau_{domain} in literature prior construction from 0.02 to 0.15 and evaluated impacts on posterior estimates and predictive performance.

## S9. Expanded human oversight log, reproducibility artifacts, and governance

### S9.1 Oversight taxonomy: operational facilitation vs scientific intervention

To reduce ambiguity about “autonomy,” we distinguish:

- **Operational facilitation:** actions required to complete execution as specified in pre-written instructions without changing scientific content (e.g., providing file paths referenced in the instructions; re-invoking an instruction step when runtime failures produced empty outputs; restarting an interrupted job).
- **Scientific intervention:** actions that change analytic content in response to intermediate results (e.g., editing concept sets; modifying cohort logic; adding/removing covariates; changing estimators; tuning model parameters; rewriting instructions to alter behavior).

Only operational facilitation occurred during agent execution. No scientific intervention was performed between steps or between runs.

### S9.2 Oversight events log

For each run (trial × run), we recorded whether any instruction step was re-invoked due to runtime errors or empty outputs, with timestamps and the affected step. When re-invocations occurred, the same instruction document and the same inputs were used. Investigators did not introduce new phenotype logic, revised eligibility criteria, or manual cohort edits during execution.

### S9.3 Artifact inventory and availability

For reproducibility, the following artifacts were retained for each run:

- instruction documents (markdown) for all six steps
- agent execution logs and intermediate reasoning traces (where available)
- generated Python scripts and executed notebooks
- OMOP concept sets (YAML/CSV) with provenance (search terms, candidate lists)
- cohort definitions and counts (CSV)
- covariate sets and balance diagnostics (SMD tables)
- model outputs (Cox/IPTW diagnostics, hazard ratios, SEs)
- discrepancy tables, quality flags, and diagnostic reports
- calibration model scripts and posterior summaries

File naming conventions and a manifest (optionally including checksums/hashes) are provided in the Supplementary Appendix.

### S9.4 Reproducibility statement for external sites

Reproduction at another institution requires: (i) OMOP CDM conformance, (ii) access to the same or equivalent drug/outcome concept sets, and (iii) execution of the instruction-driven pipeline against local data within local governance constraints. Because patient-level data cannot be shared, reproducibility is supported via sharing of instruction documents, code, and run-level artifacts. Site-specific differences in coding and care patterns are expected and are precisely the target of the proposed transport characterization.

### S9.5 Ethical considerations and LLM disclosure

This study was approved by the Institutional Review Board of the Icahn School of Medicine at Mount Sinai (protocol number to be inserted). All data remained within the HIPAA-compliant institutional environment. Use of LLM-based tools is disclosed per journal guidance. The agent model (GPT-5.2), access method (Azure OpenAI API), and access dates (May–June 2025) are reported, along with a description of the instruction documents and the boundaries of human oversight.

## Supplementary Results

## Notes

### Competing Interest Statement

K.H. and Y.Q. are co-founders of Phylo and are full-time employees of the company. B.S.G. serves as an advisor to Phylo. K.H. and Y.Q. contributed to this manuscript as co-authors; however, Phylo did not provide funding or resources for this work and, as a company, had no role in decisions regarding study design, analysis strategy, interpretation, or publication. Only the open-source version of Biomni was used. All other co-authors report no conflict of interest.

### Funding Statement

This study did not receive any funding

### Author Declarations

This study was approved by the Institutional Review Board (IRB) of the Icahn School of Medicine at Mount Sinai (STUDY-20-00338). A waiver of informed consent and HIPAA authorization was granted, as the study involved retrospective and prospective review of electronic health records with no direct patient interaction or intervention.

